# Comparison of immunogenicity between BNT162b2 and ChAdOx1 SARS-CoV-2 vaccines in a large haemodialysis population

**DOI:** 10.1101/2021.07.09.21260089

**Authors:** Candice L. Clarke, Paul Martin, Sarah Gleeson, Tina Thomson, Helena Edwards, Paige Mortimer, Stacey McIntyre, Jaid Deborah, Alison Cox, Graham Pickard, Liz Lightstone, David Thomas, Stephen P. McAdoo, Peter Kelleher, Maria Prendecki, Michelle Willicombe, in collaboration with the OCTAVE Study Consortium

## Abstract

**Background:** Limited data exists on the immunogenicity of vector-based SARS-CoV-2 vaccines in patients with kidney disease. Given their use in over 180 countries, such data is of upmost importance to inform policy on optimal vaccination strategies. This study compares the immunogenicity of BNT162b2 with ChAdOx1 in patients receiving haemodialysis.

**Methods:** 1021 patients were screened for spike protein antibodies (anti-S) following 2 doses of either BNT162b2 (n=523) or ChAdOx1 (n=498). 191 patients underwent assessment with T-cell ELISpot assays. 65 health care workers were used as a control group.

**Results:** Anti-S was detected in 936 (91.2%) of patients post-vaccination. There was no difference in seroconversion rates between infection-naïve patients who received BNT162b2, 248/281 (88.3%), compared with ChAdOx1, 227/272 (83.5%), p=0.11. Anti-S concentrations were higher following BNT162b, 462(152-1171) BAU/ml, compared with ChAdOx-1 79(20-213) BAU/ml, p<0.0001. Immunosuppression was associated with failure to seroconvert (p<0.0001); whilst being active on the transplant wait list was a predictor for seroconversion (p=0.02).

Only 73 (38.2%) of patients had detectable T-cell responses post-vaccination, with no proportional difference between infection-naïve patients who received BNT162b2, 2/19 (10.5%), versus ChAdOx1, 15/75 (20.0%), p=0.34. There were no quantitative differences in T-cell responses in infection-naïve patients, with a median 2(0-16) SFU/10^6^ PBMCs and 10(4-28) SFU/10^6^ PBMCs in those receiving BNT162b2 and ChAdOx1 respectively, p=0.35. These responses were significantly weaker compared with healthy controls.

**Conclusions:** Enhanced immunogenicity was seen with BNT162b2 compared with ChAdOx1, driven by superior humoral responses, with attenuated T-cell responses to both vaccines. Comparative data on clinical efficacy is now required.

**Significance Statement:** Limited data exist on the immunogenicity of vector-based SARS-CoV-2 vaccines in patients with kidney disease. Given their use in over 180 countries worldwide, such data are of upmost importance to inform policy on optimal vaccination strategies. This study compares the immunogenicity of BNT162b2 (n=523) against the adenovirus vector vaccine, ChAdOx1 (n=498), in 1021 haemodialysis patients. In infection-naïve patients, overall seroconversion rates were comparable, however, spike protein antibody concentrations were significantly higher following BNT162b2. No difference in T-cell responses was seen, however, all naïve patients had weaker responses compared with healthy controls. Equivalent attenuated cellular responses to both vaccines, with greater humoral responses to BNT162b2, suggests BNT162b2 has superior immunogenicity in this patient population, with data on clinical efficacy required.

---

Encouraging data on the immunogenicity of mRNA SARS-CoV-2 vaccines in haemodialysis patients has emerged globally, with seroconversion rates of >88% reported^1-4^. Whilst the kidney community awaits evidence on the corresponding clinical efficacy of mRNA vaccines, limited data exists on the immunogenicity of SARS-CoV-2 vaccines utilising alternative platforms of delivery. To date, the adenovirus vector vaccine, ChAdOx1 nCoV-19 (AZD1222), has been used in 178 countries, with 400 million doses procured by the COVID-19 Vaccine Global Access Scheme (COVAX)^5,6^. Therefore, it is prudent that data on the immunogenicity and efficacy of vector-based vaccines in patients with kidney disease are available, to help inform policy on optimal vaccine strategies in this population.

In addition, most SARS-CoV-2 immunogenicity studies in dialysis patients have focused on serological responses alone, however assessment of T-cell responses is also important when investigating and comparing adaptive immune responses to vaccination. Whilst it is recognised in healthy individuals that vector-based vaccines elicit a more robust T-cell response compared with mRNA vaccines, the accelerated immunosenescence associated with end stage kidney disease (ESKD) may have a differential effect on the cellular and humoral components of the individual vaccine responses^7,8^. Understanding these differences may drive hypotheses for optimising booster vaccines in this population, possibly using heterologous regimens.

In this study we compare the immunogenicity of the BNT162b2 and ChAdOx1 vaccine in a large, well characterised haemodialysis population, assessing both humoral and cellular components of responses and comparing them with healthy controls.

## METHODS

### Patient Selection

One-thousand, three-hundred and seventy-one patients who received haemodialysis within Imperial College Renal and Transplant Centre on or after the 8^th^ December 2020 were screened for inclusion. Patients were eligible for participation if they had received 2 doses of a SARS-CoV-2 vaccine and had a serological sample taken for analysis ≥14 days following their 2^nd^ inoculation. Of the 1371 patients screened, 1177 (85.8%) were fully vaccinated, 103 (7.5%) had declined vaccination and 91 (6.6%) were either awaiting their 2^nd^ vaccination, had missing data or had died prior to the end of the follow up period on 15^th^ June 2021 (**Figure 1**). Of the 1177 vaccinated patients, 156 (13.2%) patients did not have an eligible serological sample. Clinical and vaccine data were obtained from electronic patient records and the institutional vaccine database, respectively. The study was approved by the Health Research Authority, Research Ethics Committee (Reference: 20/WA/0123 - The Impact of COVID-19 on Patients with Renal disease and Immunosuppressed Patients).

**Figure 1.**
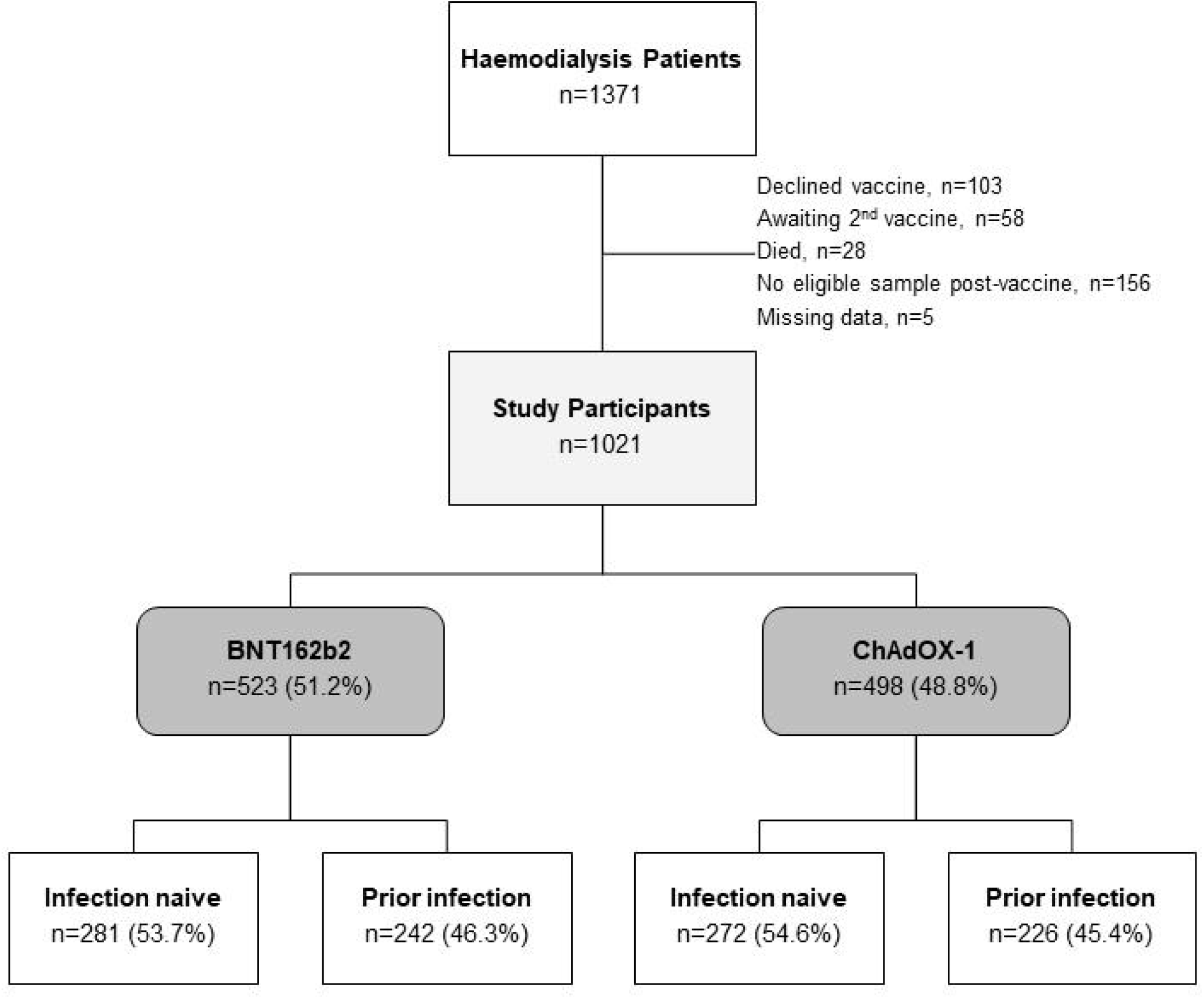
Study flow diagram.

A subgroup of 191 (18.7%) patients underwent more in-depth immunological analysis of serological and cellular responses to SARS-CoV-2 vaccination. These patients included participants of the OCTAVE study, an Observational Cohort Study of T-cells, Antibodies and Vaccine Efficacy in SARS-CoV-2 in people with chronic diseases and/or secondary immunodeficiency, which is part of the UK COVID-19 Immunity National Core Study Programme. The OCTAVE study was approved by the Health Research Authority, Research Ethics Committee (Reference:21/HRA/0489). A group of 65 healthcare workers (HCW), with a median age of 38 (30-46) years, was used as a comparator group for this in-depth analysis. Fifty and 15 HCW received the BNT162b2 and ChAdOx1 vaccines, respectively. The median interval between vaccinations in the HCW was 68 (61-70) days, with a median time to sampling post-vaccination of 28 (21-28) days.

### SARS-CoV-2 antibody detection

Serological sampling was performed during haemodialysis sessions. Serum was tested for antibodies to both the nucleocapsid protein (anti-NP) and spike protein (anti-S). Anti-NP was tested using the Abbott Architect SARS-CoV-2 IgG 2 step chemiluminescent immunoassay (CMIA) according to manufacturer’s instructions. This is a non-quantitative assay and samples were interpreted as positive or negative with a threshold index value of 1.4. The presence of anti-NP was used as a marker of natural infection. For vaccine responses, anti-S IgG were assessed using the Abbott Architect SARS-CoV-2 IgG Quant II CMIA. Anti-S antibody titres are quantitative with a threshold value of 7.1 BAU/ml for positivity, and an upper level of detection of 5680 BAU/ml.

### Detection of cellular responses to SARS-CoV-2

SARS-CoV-2 specific T-cell responses were detected using the T-SPOT® Discovery SARS-CoV-2, and assays performed by Oxford Immunotec. In brief, peripheral blood mononuclear cells (PBMCs) were isolated from whole blood samples with the addition of T-Cell Select™ (Oxford Immunotec) where indicated. 250,000 PBMCs were plated into individual wells of a T-SPOT® Discovery SARS-CoV-2 plate. The assay measures immune responses to SARS-CoV-2 spike protein peptide pools (S1 protein and S2 protein), in addition to positive PHA (phytohemagglutinin) and negative controls. Cells were incubated and interferon-γ secreting T cells were detected. Spot forming units (SFU) were detected using an automated plate reader (Autoimmun Diagnostika). Infection-naïve, unvaccinated participants were used to identify a threshold for a positive response using mean +3 standard deviation SFU/10^6^ PBMC, as previously described^9^. This resulted in a cut-off for positivity of 40 SFU/10^6^ PBMC, established by Imperial College London/North-West London Pathology.

### Definition of prior SARS-CoV-2 infection

Prior exposure was defined by a history of infection confirmed through viral detection from nasopharyngeal swab specimens, via reverse-transcriptase polymerase chain reaction (RT-PCR) assays, or by serological assessment. Routine symptomatic screening of patients occurred prior to each haemodialysis session from the 9^th^ March 2020 onwards. In addition, routine asymptomatic swabbing was performed in June 2020, and recommenced weekly from 15^th^ November 2020 onwards. Protocolised serological screening of haemodialysis patients had commenced in our centre in June 2020, and these data were used to aid identification of patients with prior asymptomatic SARS-CoV-2 exposure prior to vaccination. As part of this protocol, patients were initially screened for anti-NP, and those with a subthreshold anti-NP index value (0.25-1.4), underwent confirmatory testing for natural infection by assessing for receptor binding domain (anti-RBD) antibodies. This was performed using an in-house double binding antigen ELISA (Imperial Hybrid DABA; Imperial College London, London, UK), which detects total RBD antibodies^10-13^

### Statistical Analysis

Statistical analysis was conducted using Prism 9.0 (GraphPad Software Inc., San Diego, California). Unless otherwise stated, all data are reported as median with interquartile range (IQR). The Chi-squared test was used for proportional assessments. The Mann-Whitney and Kruskal-Wallis tests were used to assess the difference between 2 or >2 groups, with Dunn’s post-hoc test to compare individual groups. Multivariable analysis was carried out using logistic regression using variables which were found to be significant on univariable analysis.

## RESULTS

1021 patients were tested for anti-S, at a median time of 41 (27-49) days following their 2^nd^ SARS-CoV-2 vaccine. Of these patients, 465/1021 (45.8%) had evidence of natural infection; 341 (72.9%) having been diagnosed via RT-PCR testing, with an additional 127 (27.1%) having a diagnosis made serologically. 523 (51.2%) patients received the BNT162b2 vaccine and 498 (48.8%) received the ChAdOx1 vaccine (**Figure 1**). There was no difference in the proportion of patients with prior infection between those receiving BNT162b2, 242 (45.3%), compared with ChAdOx1, 226 (45.4%), p=0.87.

### Serological responses in infection-naïve patients

The overall seroconversion rate in infection-naïve patients was 475/553 (85.9%). There was no difference in seroconversion rates between infection-naïve patients who received BNT162b2, 248/281 (88.3%) patients, compared with 227/272 (83.5%) patients who received ChAdOx1, p=0.11. However, anti-S concentrations were significantly higher in patients who received BNT162b, with median anti-S concentrations in the BNT162b and ChAdOx1 patients of 462 (152-1171) BAU/ml and 79 (20-213) BAU/ml respectively, p<0.0001 (**Figure 2a**).

**Figure 2.**
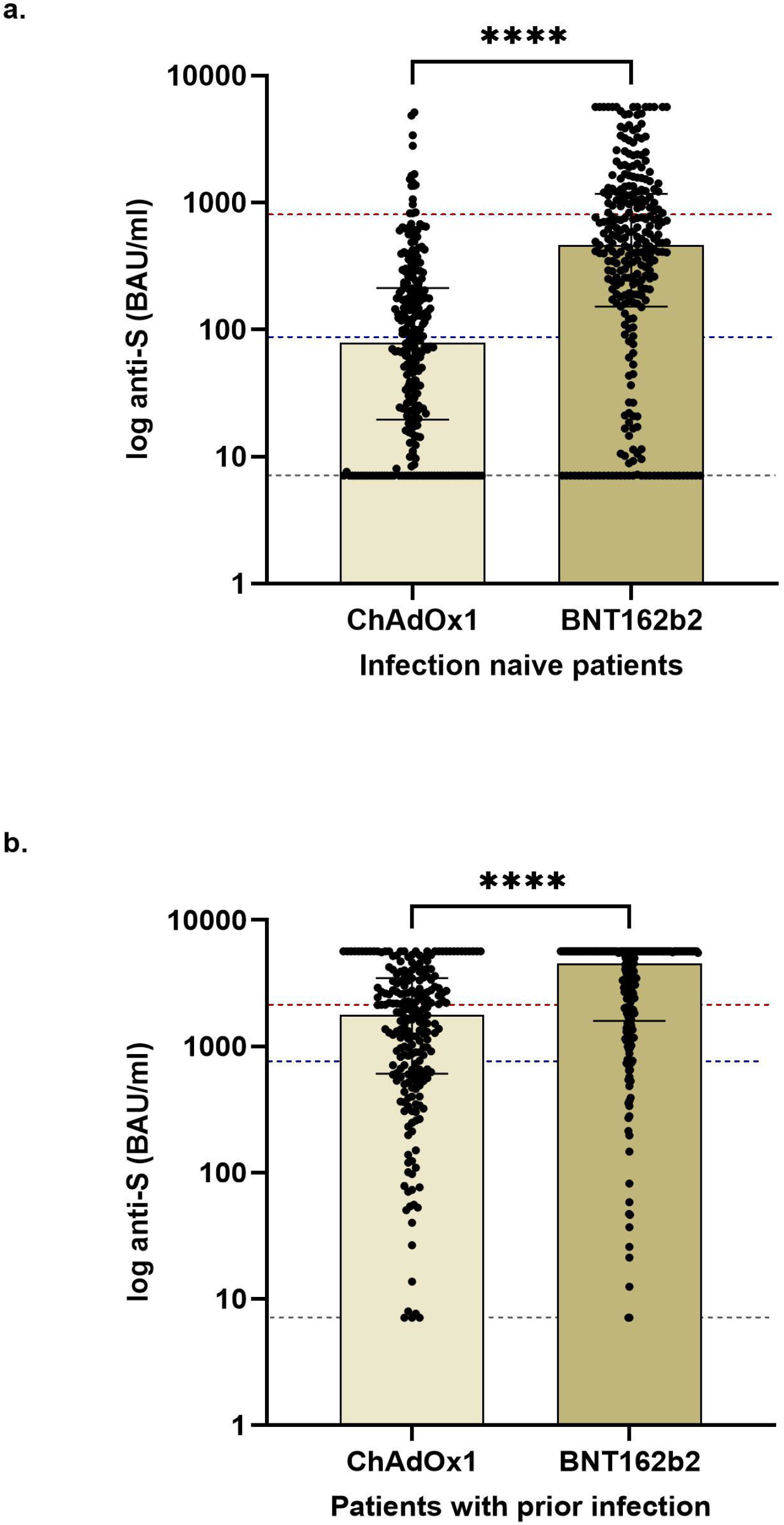
Comparison of spike protein antibodies in haemodialysis patients receiving BNT162b2 compared with ChAdOx1, **a. In infection-naïve patients**. The median anti-S concentrations in the BNT162b and ChAdOx1 patients were 462 (152-1171) and 79 (20-213) BAU/ml respectively, p<0.0001. The dotted lines represents the median anti-S of infection-naive health care workers (HCW) who received the BNT162b2 vaccine, 815 (318-2033) BAU/ml (red line) and ChAdOx1 vaccine, 88 (47-395) BAU/ml (blue line). The black dotted line indicates the positive cut off of the assay 7.1 BAU/ml (black line). **b. In patients with prior infection**. The median anti-S concentrations in the BNT162b and ChAdOx1 patients were 4523 (1593-5680) and 1788 (608-3477) BAU/ml respectively, p<0.0001. The dotted lines represents the median anti-S of HCW with prior infection who received the BNT162b2 vaccine, 2189 (1236-3303) BAU/ml (red line) and ChAdOx1 vaccine, 753 (574-867) BAU/ml (blue line).

Clinical characteristics associated with a reduced likelihood of seroconverting included length of time at end stage kidney disease (ESKD), the concomitant use of immunosuppression and having received a previous transplant (**Table 1**). Being active on the transplant wait list was associated with increased seroconversion rates (**Table 1**). Comparison of anti-S concentrations demonstrated lower levels in patients receiving immunosuppression compared with non-immunosuppressed patients, with median concentrations of 27 (7.1-338) BAU/ml and 231 (53-364) BAU/ml respectively, p<0.0001 (**Figure 3a**). Analysing concentrations by immunosuppression type showed that patients receiving calcineurin inhibitor (CNI) monotherapy, with a median anti-S concentration of 31 (7.1-154) BAU/ml, had significantly lower concentrations compared with those receiving prednisolone monotherapy, 191 (18-1335) BAU/ml, p=0.028; but comparable concentrations to those patients on combination immunosuppression, who had a median anti-S concentration of 8 (7.1-198) BAU/ml, p=0.47 (**Figure 3b**). Patients who had received a transplant previously also had lower median anti-S concentrations compared with those who had not at 121 (7.1-428) BAU/ml and 209 (37-679) BAU/ml respectively, p=0.002 (**Figure 3c**). In previously transplanted patients, anti-S concentrations were significantly lower in those still receiving immunotherapy, 26 (7.1-163) BAU/ml, compared with those in whom immunosuppression had been withdrawn, 397 (171-885) BAU/ml), p<0.0001 (**Figure 3d**). There were no quantitative differences in anti-S concentrations according to gender, ethnicity, cause of ESKD or a diagnosis of diabetes (*Supplemental Information* **Figure S1**). No correlation was seen between age and serological responses, p=0.28.

**Table 1.**
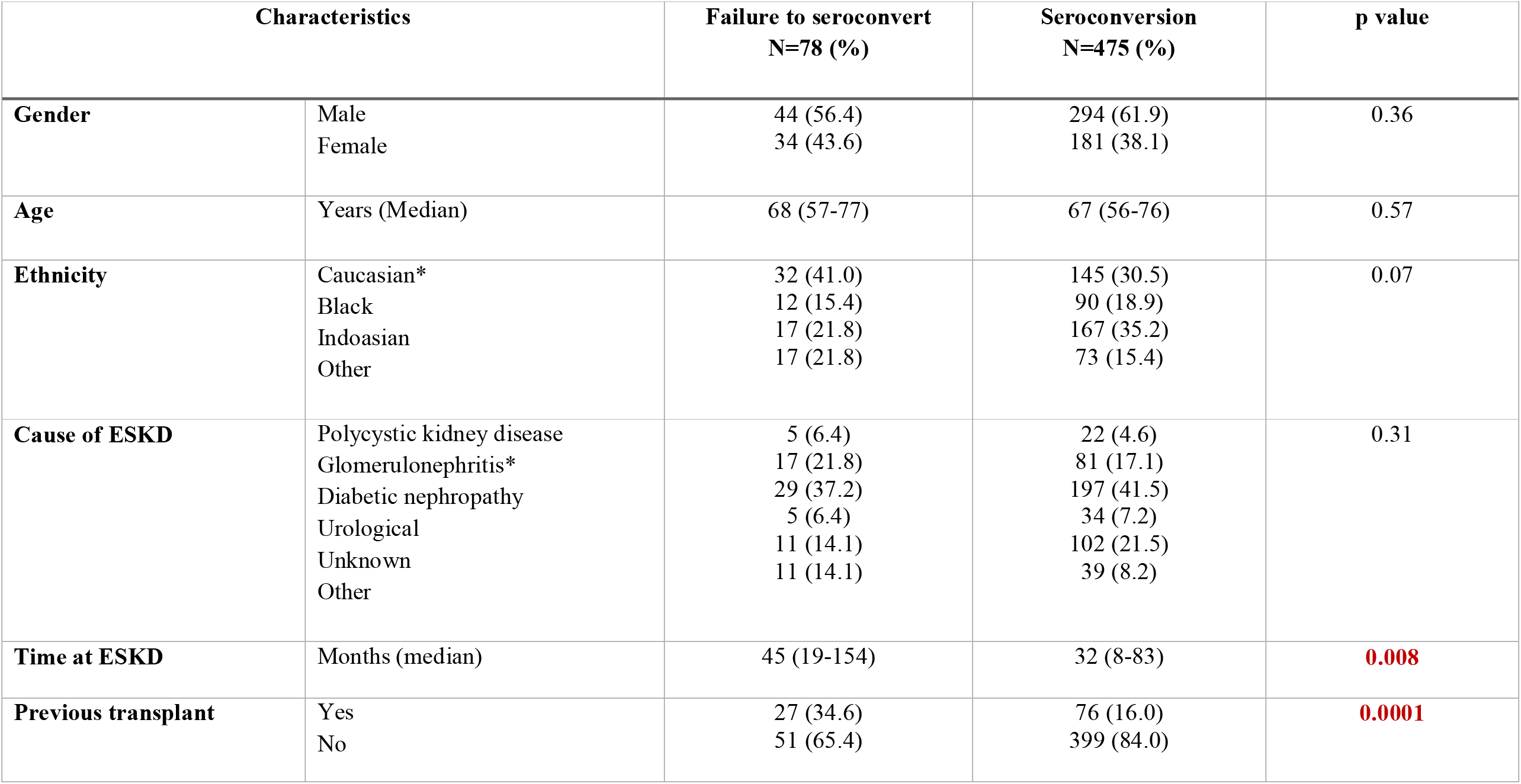
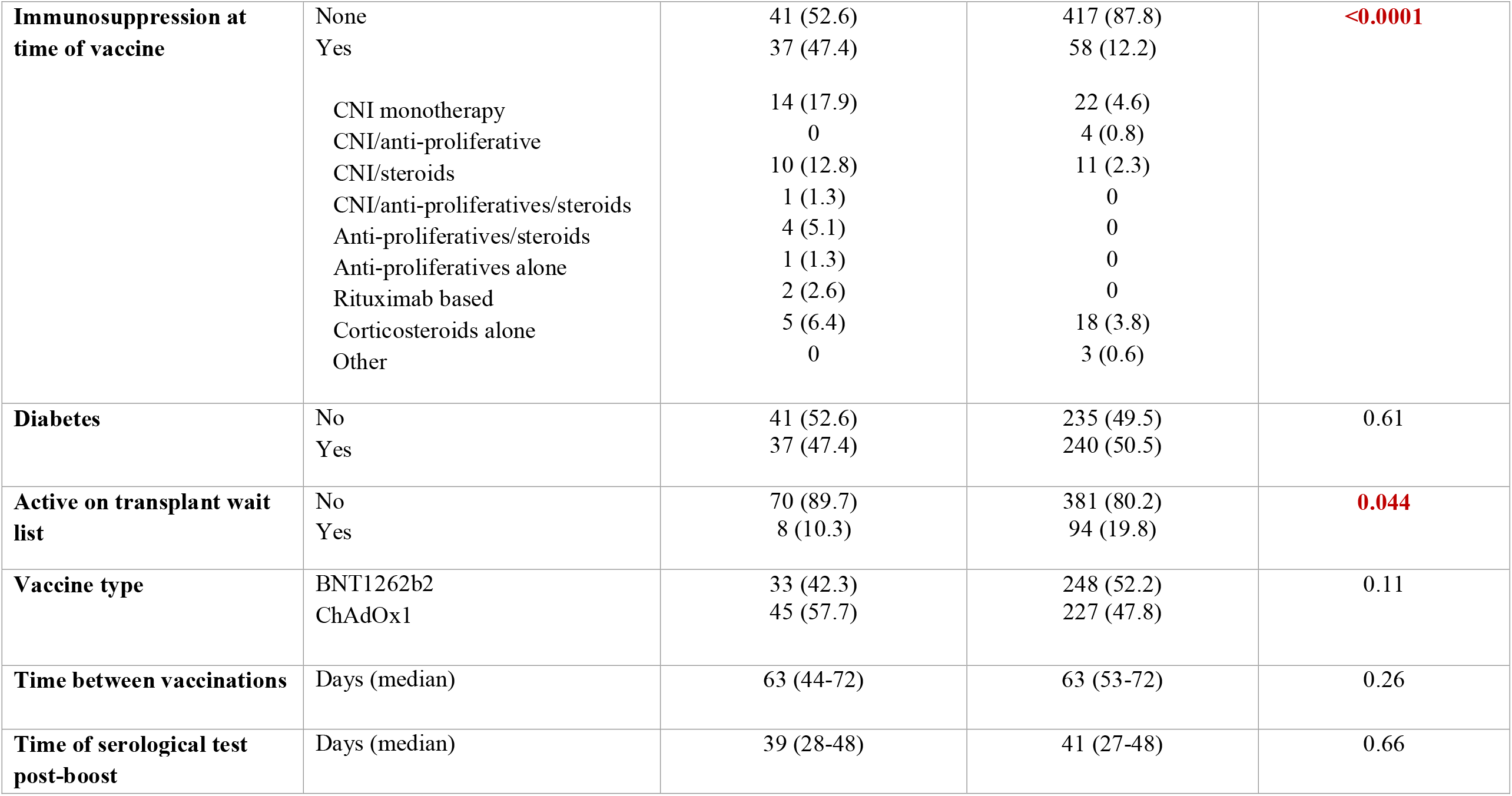
Clinical characteristics associated with seroconversion following SARS-CoV-2 vaccination in infection naïve haemodialysis patients.

**Figure 3.**
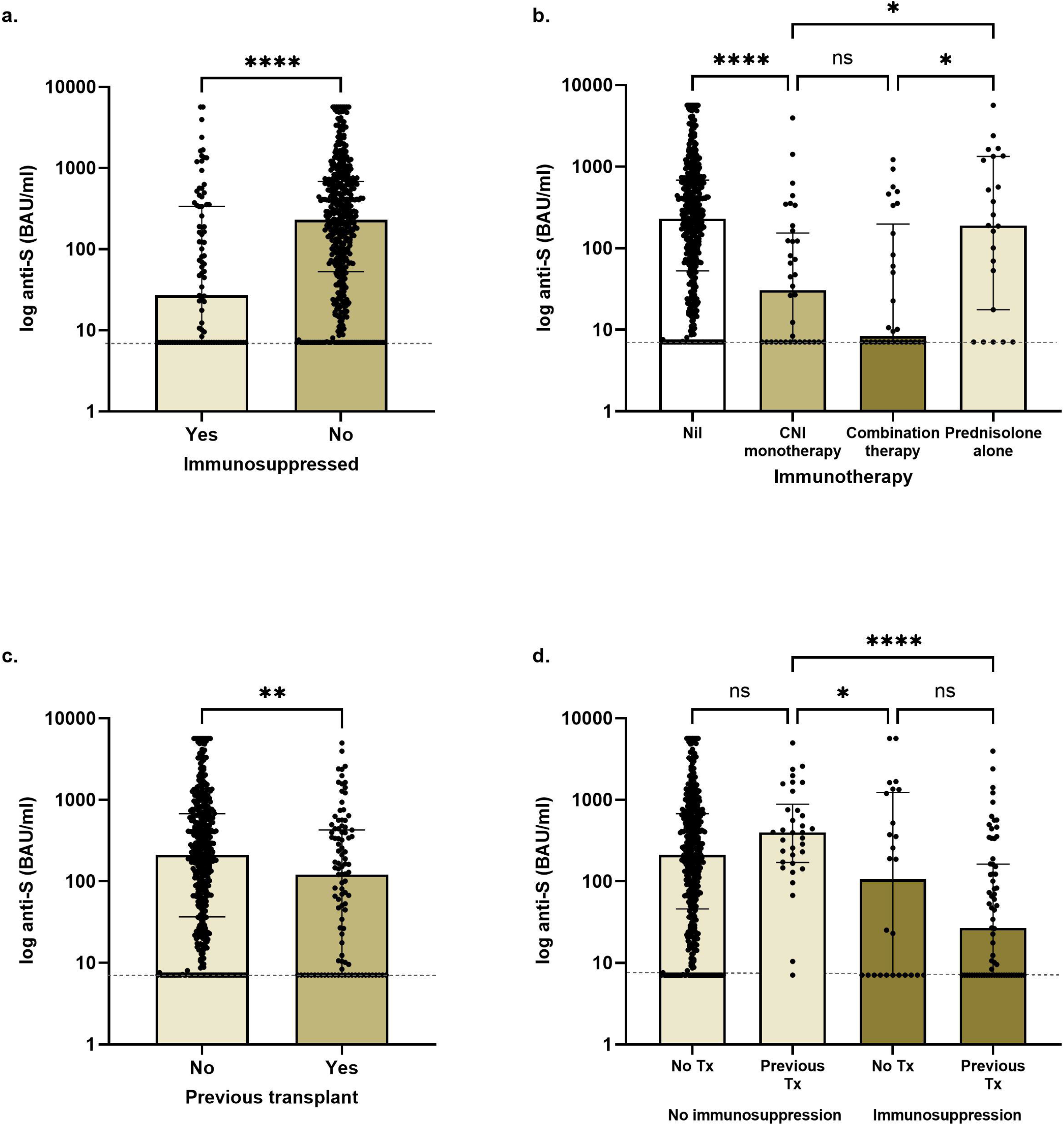
Comparison of spike protein antibodies in infection-naïve haemodialysis patients by immunosuppression history. a. Patients receiving immunosuppression had significantly lower anti-S concentrations, 27 (7.1-338) BAU/ml compared with patients not receiving maintenance immunosuppression, 231 (53-684) BAU/ml, p<0.0001. b. Median anti-S concentrations in patients receiving CNI monotherapy, combination therapy or prednisolone monotherapy were 31 (7.1-154) BAU/ml, 8 (7.1-198) BAU/ml and 191 (18-1335) BAU/ml, respectively. With significant differences between patients receiving CNI monotherapy and prednisolone, p=0.028 but no difference between patients receiving CNI monotherapy compared with combination therapy, p=0.47. c. Patients who had previously had a transplant had lower anti-S concentrations, 121 (7.1-428) BAU/ml compared with patients who had never received a transplant, 209 (37-688), p=0.002. d. For previously transplanted patients, anti-S concentrations were significantly higher in those patients in whom immunosuppression had been withdrawn, 397 (171-885) BAU/ml), compared with those patients still receiving immunosuppression, 26 (7.1-163) BAU/ml p<0.0001.

On multivariable analysis, use of immunosuppression, OR 0.15 (0.09-0.26), p<0.0001 was associated with non-seroconversion; whilst being active on the transplant wait list remained an independent predictor of seroconversion, OR 2.61 (1.16-5.87), p=0.02 (*Supplemental Information* **Table S1**).

### Serological responses in patients with prior infection

Of 468 patients with prior infection, 5 (1.1%) remained seronegative post-vaccination; 2/226 (0.9%) who received ChAdOx1 and 3/242 (1.2%) who received BNT162b2, p=0.71. All 5 patients had prior infection diagnosed by PCR testing, and the individual patient characteristics maybe found in the *Supplemental Information* (**Table S2**). The overall median anti-S concentrations post vaccination were higher in patients with prior exposure compared with infection-naïve patients, at 2702 (980-5680) BAU/ml and 189 (27-628) BAU/ml respectively, p<0.0001.

Considering patients with prior infection independently, those patients who received ChAdOx1 had a median anti-S of 1788 (608-3477) BAU/ml, which was significantly less than patients who had received BNT162b2, who had a median anti-S of 4523 (1593-5680) BAU/ml, p<0.0001 (**Figure 2b**). There was no difference in median anti-S in those patients who were diagnosed by PCR compared with serology, with patients receiving BNT162b2 having a median anti-S of 5184 (1773-5680) BAU/ml and 3221 (1379-5680)) BAU/ml respectively, p=0.70, and those receiving ChAdOx1 having median concentrations of 1183 (512-2807) BAU/ml and 2080 (675-3876) BAU/ml respectively, p=0.21 (*Supplemental Information* **Figure S2**). There was a negative correlation between the anti-NP index and time of test post PCR confirmed infection, r=-0.30, p<0.0001. The co-existence of anti-NP on post-vaccination testing was associated with higher anti-S concentrations, with anti-NP+ and anti-NP-patients receiving BNT162b2 having a median anti-S of 5680 (3338-5680) BAU/ml and 2802 (1097-5680) BAU/ml respectively, p<0.0001; and those receiving ChAdOx1, 2120 (880-4485) BAU/ml and 1472 (636-273) BAU/ml respectively, p=0.01 (*Supplemental Information* **Figure S2)**.

### Cellular and humoral responses in infection-naïve patients

A subgroup of 191 patients had paired assessment of T-cell and serological responses at a median time of 27 (26-28) days post-vaccination. Fifty of 191 (26.2%) patients received BNT162b2 and 141 (73.8%) received ChAdOx1. Nineteen of 50 (38.0%) patients who received BNT162b2 and 75 (53.1%) of those who received the ChAdOx1 vaccines were infection-naïve, p=0.052.

Overall, 17/94 (18.1%) of infection-naïve patients had detectable T-cell responses post-vaccination. There was no proportional difference in ELISpot positivity in infection-naïve patients who received BNT162b2, 2/19 (10.5%), compared with ChAdOx1, 15/75 (20.0%), p=0.34. On quantification of cellular responses, there was no difference in the median number of spot forming units (SFU/10^6^ PBMCs) between infection-naïve patients who received BNT162b2 compared with ChAdOx1, at 2 (0-16) SFU/10^6^ PBMCs and 10 (4-28) SFU/10^6^ PBMCs respectively, p=0.35. However, compared with infection-naïve HCW receiving the corresponding vaccine, responses were significantly weaker in patients, with infection-naïve HCW receiving BNT162b2 and ChAdOx1 having a median 63 (21-132) SFU/10^6^ PBMCs, p<0.0001 and 68 (30-162) SFU/10^6^ PBMCs, p=0.0083 respectively (**Figure 4a**). Serological assessment of these infection-naïve patients demonstrated no difference in seroconversion rates in patients following vaccination with either BNT162b2 or ChAdOx1, with 14/19 (73.7%) and 57/75 (76.0%) patients seroconverting respectively, p=0.83. Quantification of these responses showed significantly higher anti-S concentrations in patients who received BNT162b2 compared with ChAdOX1, with anti-S concentrations of 557 (7.1-1745) BAU/ml and 82 (8-183) BAU/ml respectively, p=0.02 (**Figure 4b**). However, there were no differences in anti-S concentrations between infection-naïve HCW and patients who received the corresponding vaccine, with HCW receiving BNT162b2 having a median anti-S of 815 (318-2033), p=0.054, and those receiving ChAdOx1 a median anti-S of 88 (47-395) BAU/ml, p=0.97 (**Figure 4b**).

**Figure 4.**
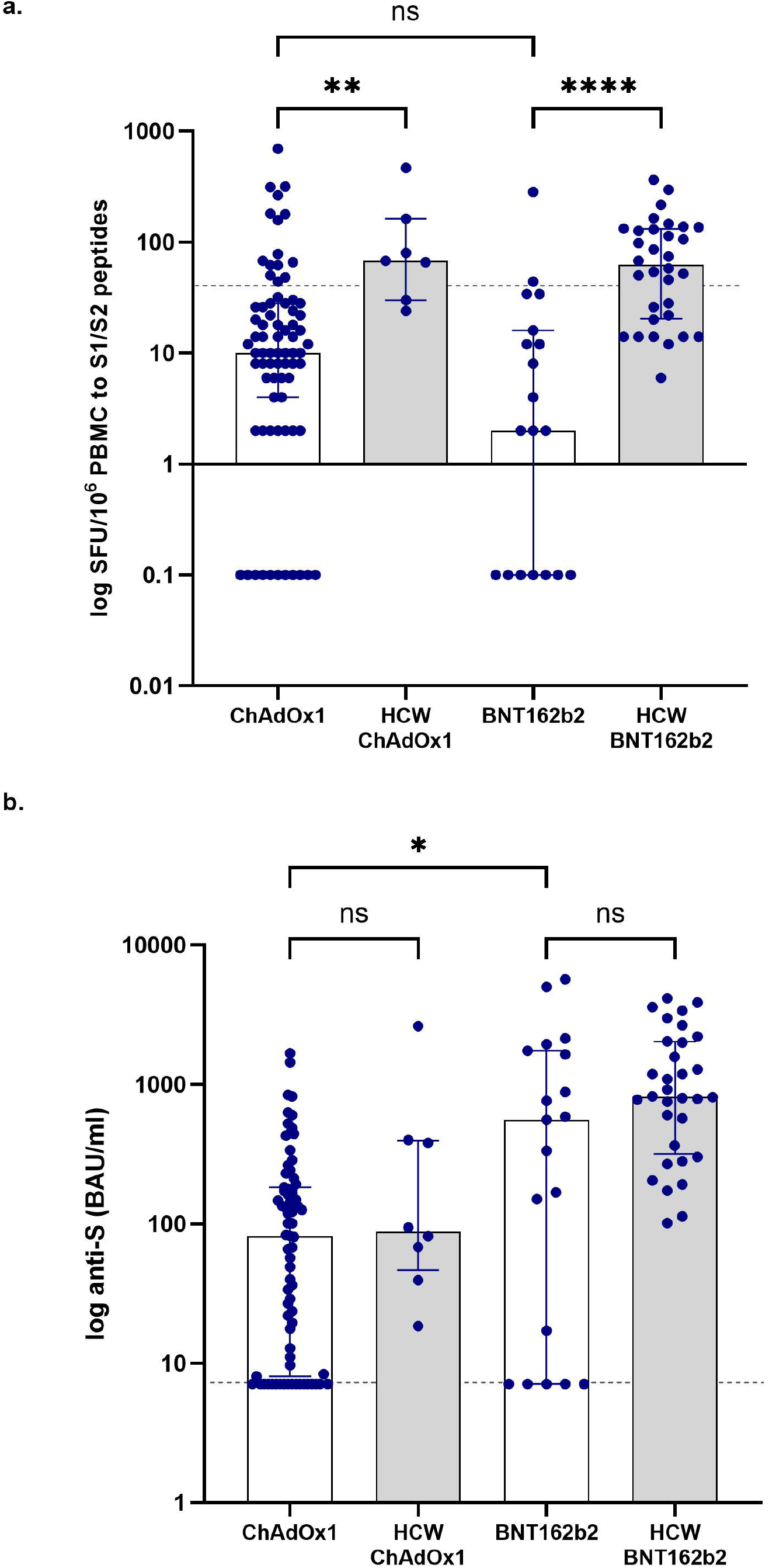
Comparison of post-vaccination T-cell and serological responses in infection-naïve patients and healthcare workers. The median SFU in infection-naïve patients receiving ChAdOx1 and BNT162b2 were 10 (4-28) and 2 (0-16) SFU/10^6^ PMBCs respectively. The median SFU in infection-naïve HCW receiving ChAdOx1 and BNT162b2 was 68 (30-162) SFU/10^6^ and 63 (20.5-131.5) SFU/10^6^ PMBCs respectively. The median anti-S concentrations in infection-naïve patients receiving ChAdOx1 and BNT162b2 were 82 (8.0-183) and 557 (7.1-1745) respectively. The median anti-S concentrations in infection-naïve HCW receiving ChAdOx1 and BNT162b2 were 88 (47-395) and 815 (318-2033) BAU/ml respectively. a. There was no difference in SFU in patients receiving ChAdOx1 compared with BNT162b2, p=0.35. Responses were significantly weaker in patients compared with HCW receiving the same vaccine; ChAdOx1, p=0.0083 and BNT162b2, p<0.0001. b. There was no difference in anti-S concentrations in patients compared with HCW receiving the same vaccine; ChAdOx1, p=0.97 and BNT162b2, p=0.054. Patients receiving BNT162b2 had significantly higher anti-S than those receiving ChAdOx1, p=0.02.

### Cellular and humoral responses in patients with prior infection

Overall, 56/97 (57.7%) of patients with prior exposure had detectable T-cell responses post-vaccination. There was no proportional difference in ELISpot positivity in patients with prior infection who received BNT162b2, 15/31 (48.4%), compared with ChAdOx1, 41/66 (62.1%), p=0.20. On quantification of cellular responses, there was no difference in median SFU/10^6^ PBMCs in previously exposed patients who received BNT162b2 compared with ChAdOX1, with 26 (6-144) SFU/10^6^ PBMCs and 102 (18-250) SFU/10^6^ PBMCs respectively, p=0.08 (**Figure 5a**). No significant differences were seen between previously exposed HCW receiving ChAdOx1, 114 (74-216) SFU/10^6^ PBMCs, compared with patients who received ChAdOx1, p=0.99. However, previously exposed HCW who received BNT162b2 had significantly greater responses, 246 (131-332) SFU/10^6^ PBMCs, compared with patients receiving BNT162b2, p=0.0017 (**Figure 5a**).

**Figure 5.**
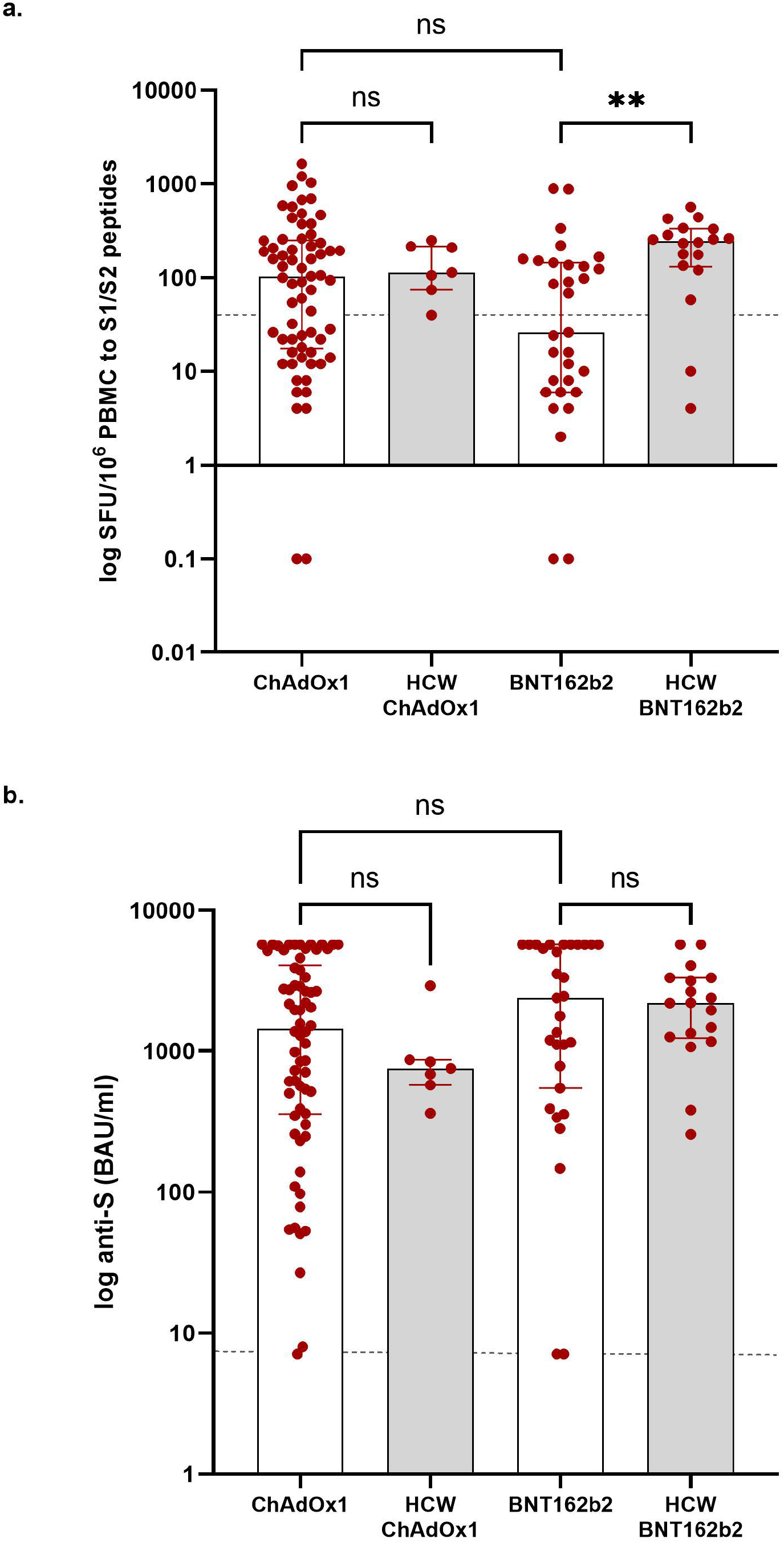
Comparison of post-vaccination T-cell and serological responses in patients and healthcare workers with prior exposure to SARS-CoV-2. The median SFU in previously exposed patients receiving ChAdOx1 and BNT162b2 were 102 (18-250) and 26 (6-144) SFU/10^6^ PMBCs respectively. The median SFU in previously exposed HCW receiving ChAdOx1 and BNT162b2 were 114 (74-216) and 246 (131-332) SFU/10^6^ PMBCs respectively. The median anti-S concentrations in previously exposed patients receiving ChAdOx1 and BNT162b2 were 1446 (357-4063) and 2380 (544-5680) respectively. The median anti-S in previously infected healthcare workers receiving ChAdOx1 and BNT162b2 were 753 (574-867) and 2189 (1236-3303) BAU/ml respectively. a. There was no difference in SFU in patients receiving ChAdOx1 compared with BNT162b2, p=0.08. Responses were significantly weaker in patients compared with HCW receiving BNT162b2, p=0.0017, but not ChAdOx1, p=0.99. b. There was no difference in anti-S concentrations in patients receiving ChAdOx1 compared with BNT162b2, p=0.45. There was no difference in anti-S concentrations in patients compared with HCW receiving the same vaccine; ChAdOx1, p=0.99 and BNT162b2, p=0.92.

Ninety-four of 97 (96.9%) patients with prior evidence of infection had detectable anti-S. There was no difference in anti-S concentrations in patients with prior infection who received BNT162b2, 2380 (544-5680) BAU/ml, compared with ChAdOx1, 1446 (357-4063) BAU/ml, p=0.45. There were also no differences between patients and HCW with prior exposure who received BNT162b2, median anti-S 2189 (1236-3303) BAU/ml), p=0.99, or HCW who received ChAdOx1, median anti-S 753 (574-867) BAU/ml, p=0.92 (**Figure 5b**).

Within the 191 patients assessed, a positive correlation between serological (anti-S) and cellular responses (SFU/10^6^ PBMCs) was seen, r=0.55, p<0.0001 (**Figure 6**). Overall, 25/191 (17.7%) patients within the subgroup had no detectable immunological response, representing 23/94 (24.5%) of infection-naïve patients and 2/97 (2.1%) of patients with prior exposure. Concomitant use of immunosuppression was significantly associated with failure to mount an immunological responses, with 12/23 (52.2%) of the infection naïve patients without detectable responses receiving long-term immunosuppression (**Table 2**). A breakdown of the individual patient characteristics in those failing to make a detectable response to either vaccine is provided in the supplemental information (**Table S2**).

**Table 2.**
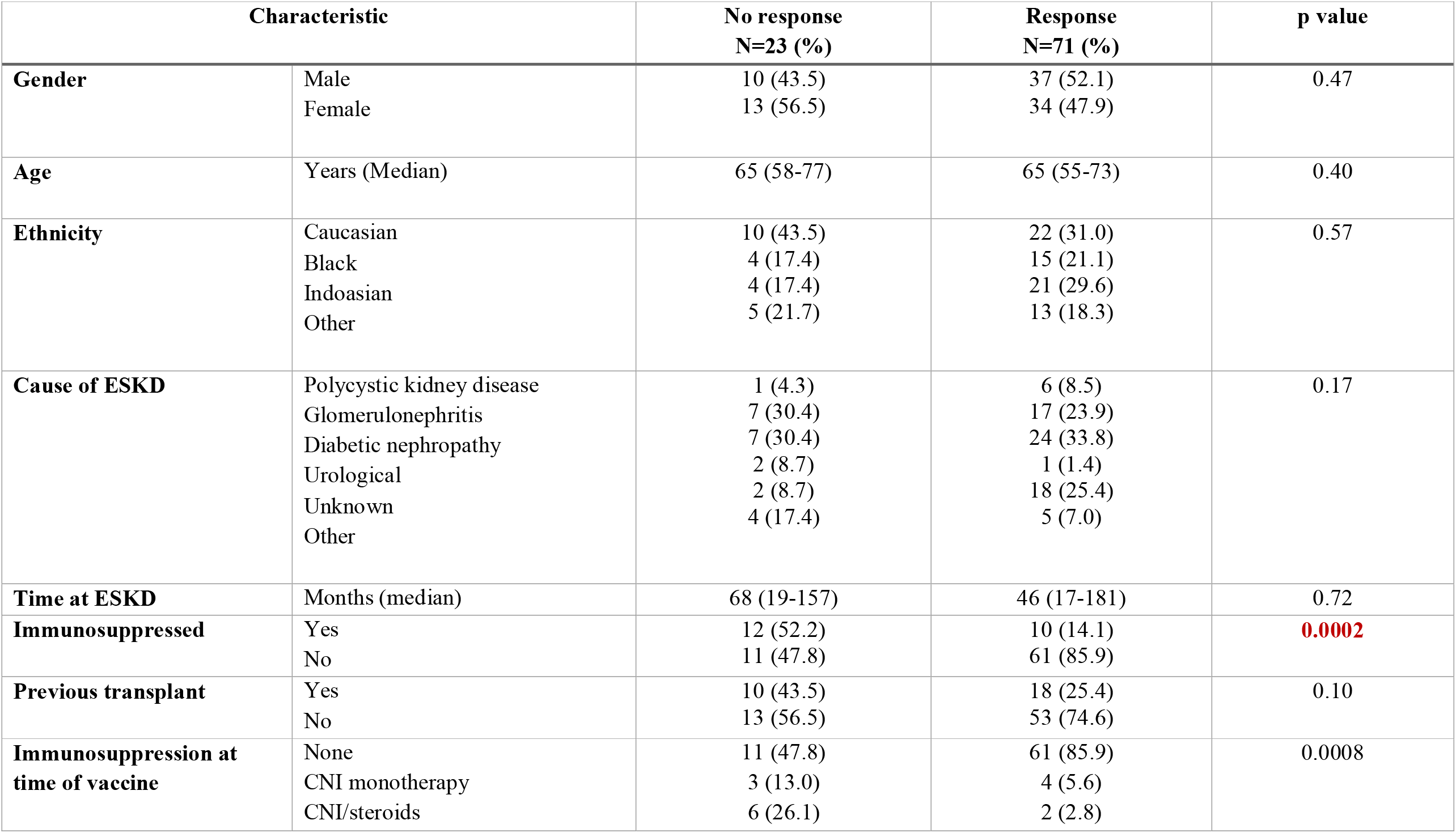
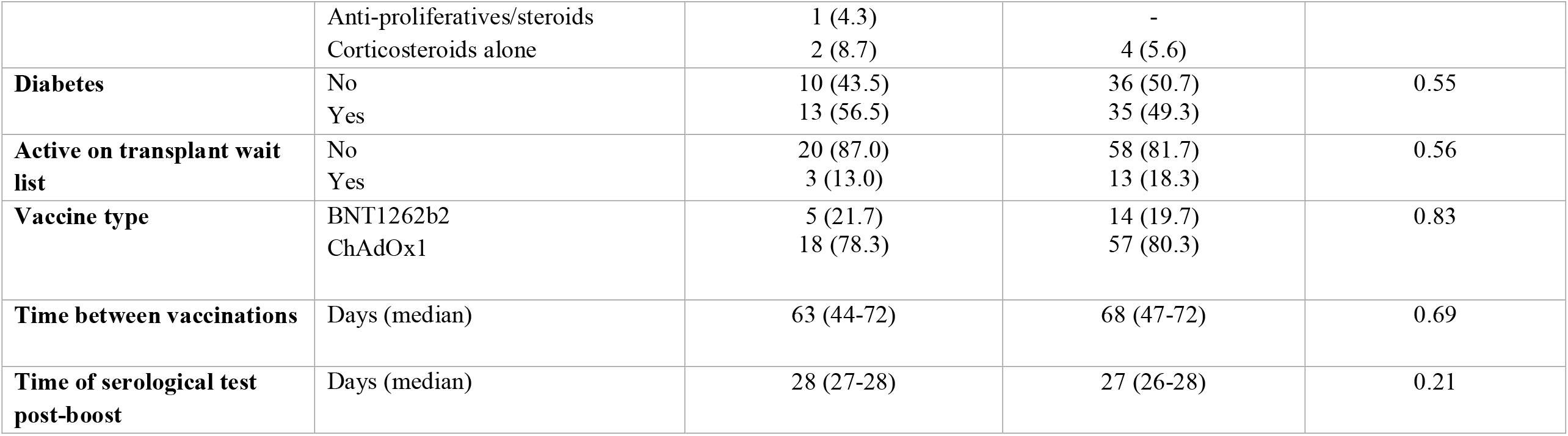
Clinical associations of non-immunological (cellular and serological) response in infection naïve patients.

## DISCUSSION

The high number of patients, 45.8%, who were found to have prior natural infection in our cohort reinforces the need for optimal vaccination strategies in this at-risk population. Overall, 936 (91.7%) of patients had detectable anti-S post-vaccination, with seroconversion occurring in 475 (85.9%) of infection-naïve patients. Whilst we found no difference in the proportion of patients seroconverting between BNT162b2 and ChAdOx1, quantitatively there were significantly higher anti-S concentrations in patients with prior exposure and infection-naïve patients, who had received BNT162b2. Whilst we do not report on neutralisation properties, which are the closest laboratory correlate to immune protection, it may be considered that anti-S IgG titres correlate with neutralising ability^14-17^. Extrapolating from this observation, one may hypothesise that dialysis patients are more likely to produce neutralising antibodies following vaccination with BNT162b2 compared with ChAdOx1.

Immunogenicity data from the vaccine clinical trials suggest that T-cell responses are more robust following ChAdOx1 compared with BNT162b2^7,18,19^. Although we found no difference in T-cell responses between the vaccines in infection-naïve patients, we observed overall blunted responses compared with the corresponding healthy controls. Conversely, in patients with previous exposure, there were no statistical differences in T-cell responses between the vaccines, but patients who had received BNT162b2 had weaker responses compared to the corresponding healthy controls. Only two other studies have reported cellular responses to mRNA vaccines in dialysis patients; both report better T-cell responses compared with our patients who received BNT162b2^20,21^. This may in part reflect the peptide pools used in the ELISpot assays or differing thresholds used to define positivity. However, it is recognised that ESKD and uraemia are associated with T-cell exhaustion and suppression of IFN-γ production, and the attenuated responses we found are in keeping with this^8,22,23^. Most importantly, although assays were processed in separate laboratories, we have used the same method which enables comparison of the different vaccine responses in patients and healthy controls.

We found that concomitant use of immunosuppression was a risk factor for lack of serological and cellular responses. Other studies have reported a similar impact on serological responses in patients receiving immunosuppression^24,25^. We have been able to further delineate that type of immunosuppression being received at the time of vaccination influences serological response, rather than immunosuppressive history. A history of prior transplantation only affects anti-S concentrations if patients were still actively receiving immunosuppression. We also found that the use of calcineurin inhibitors had a greater impact on serological responses compared with those receiving prednisolone monotherapy. Tacrolimus, which was the most commonly used immunosuppressive agent in our cohort, exerts its action on T cell responses and indirectly on B cell growth and antibody production. Tacrolimus selectively inhibits follicular CD4 T cell counts and function which may be relevant in this study given the importance of T cell dependent antibody responses in generating high affinity memory humoral response^26,27^. We did not see an effect of age on immune responses; however, we did find that being active on the transplant waitlist was independently associated with seroconversion in infection-naïve patients. We hypothesise that within our population immunosenescence may be more closely related to ‘frailty’ than age per se^28^. Although we have not provided direct evidence for this, it may be considered that patients active on the transplant wait list are well, whilst age-matched individuals not being considered for transplantation are more likely to be frail in the majority of cases.

Limitations of our study include the unknown effect of the delayed prime-boost strategy of 8-12 weeks in the UK on the immunogenicity results in the patients who received BNT162b2. However, our seroconversion rates of 88.3% are comparable with other studies who report following a 3-4 week dosing interval^1,2^. Our control group is not matched for age, and being younger there may be an overestimate of the immune responses in healthy controls^9^. In addition, our control group, especially those receiving ChAdOx1, is relatively small and therefore susceptible to type II error in reporting. However, the major strength of our study is that we are the first to comprehensively report on the immunological response to the ChAdOx1 vaccine in patients with kidney disease. This is the largest analysis of both humoral and cellular responses to date in an ESKD population, and we report detailed patient level data. Moreover, our ethnically diverse cohort has been characterised in depth throughout the pandemic with regular screening via PCR and serological testing, enabling accurate identification of individuals with prior exposure^13,29,30^.

It is important to highlight, that although we have dissected the immunogenic properties of both the BNT162b2 and ChAdOx1 vaccines to better compare their properties in a haemodialysis population, immune responses to both vaccines were encouraging in light of the recognised attenuated responses to other commonly used vaccines such as Influenza and Hepatitis B^31,32^. Whilst comparative data on their clinical effectiveness in patients receiving dialysis are awaited, we advise all patients to get vaccinated with the first vaccine which is available to them. Recent studies suggest excellent efficacy of both BNT126b2 and ChAdOx1 vaccines in preventing infections and hospitalisations in the general population, which is likely to be the case for the majority of patients with kidney disease too^33,34^. It is very likely that as the pandemic continues, and variants emerge, that everyone will require booster vaccines, which may be optimised by heterologous vaccine schedules, and data on the efficacy of different vaccines in kidney patients may aid decision making. The kidney community does need to reinforce the need to prioritise our patients for preventative strategies, which may require bespoke planning from evidence delivered from within this population with individualised complexities.

## Supporting information

Strobe

## Data Availability

Data is not openly available on an external database. Requests for information can be made to the corresponding author.

## ACKNOWLEDGEMENTS

The OCTAVE trial, which is part of the COVID-19 Immunity National Core Study Programme, was sponsored by the University of Birmingham and funded by a grant from UK Research and Innovation (UKRI) administered by the Medical Research Council (grant reference number MC_PC_20031). It has been designated an Urgent Public Health (UPH) study by the National Institute of Health Research.

This research is also supported by the National Institute for Health Research (NIHR) Biomedical Research Centre based at Imperial College Healthcare NHS Trust and Imperial College London. The authors would like to thank the West London Kidney Patient Association, all the patients and staff at ICHNT (The Imperial COVID vaccine group and dialysis staff, and staff within the North West London Pathology laboratories). The authors are also grateful for the support from Hari and Rachna Murgai, The Nan Diamond Fund and the Auchi Charitable Foundation.

