## Supplemental Information for "Comparison of immunogenicity between BNT162b2 and ChAdOx1 SARS-CoV-2 vaccines in a large haemodialysis population"

**CONTENTS**

**Figure S1. Spike protein antibody concentrations in infection-naïve patients.**

**Table S1. Multivariable analysis of factors associated with a serological response to SARS-CoV-2 vaccination in haemodialysis patients.**

**Table S2**. **Characteristics of in patients with prior infection who failed to seroconvert.**

**Figure S2. Comparison of anti-S concentrations by method of diagnosis of previous infection and the presence of co-existing anti-NP.**

**Table S3**. **Characteristics of 25 patients with no immunological response**

**Figure S1. Spike protein antibody concentrations in infection-naïve patients.**

a. Anti-S concentrations in infection-naïve male and female were 171 (28-593) and 231 (27-731) BAU/ml respectively, p=0.32.
b. There was no difference in anti-S concentrations between patients of white ethnicity compared with patients from Black backgrounds, at 147 (11-448) and 192 (34-1022) BAU/ml respectively, p=0.10. Patients from indoasian backgrounds had higher anti-S concentrations at 223 (48-639), p=0.01, compared with White patients.
c. There was no impact of cause of end stage kidney disease (ESKD) and anti-S concentrations, p=0.71; with concentrations of 163 (18-677), 121 (21-428), 202 (36-653) and 235 (22-589) BAU/ml in patients with glomerulonephritis (GN), polycystic kidneys (APKD), diabetes (DM) and urological causes of ESKD respectively.
d. There was no difference in anti-S concentrations between those patients active on the transplant wait list or not, at 176 (24-625) and 236 (73-656), p=0.20, respectively.
e. There was no difference in anti-S concentrations between those patients with or without diabetes, at 205 (33-636) and 175 (24-589) BAU/ml, p=0.63, respectively.
f. There was no correlation between age and anti-S concentrations, r=-0.05, p=0.28.

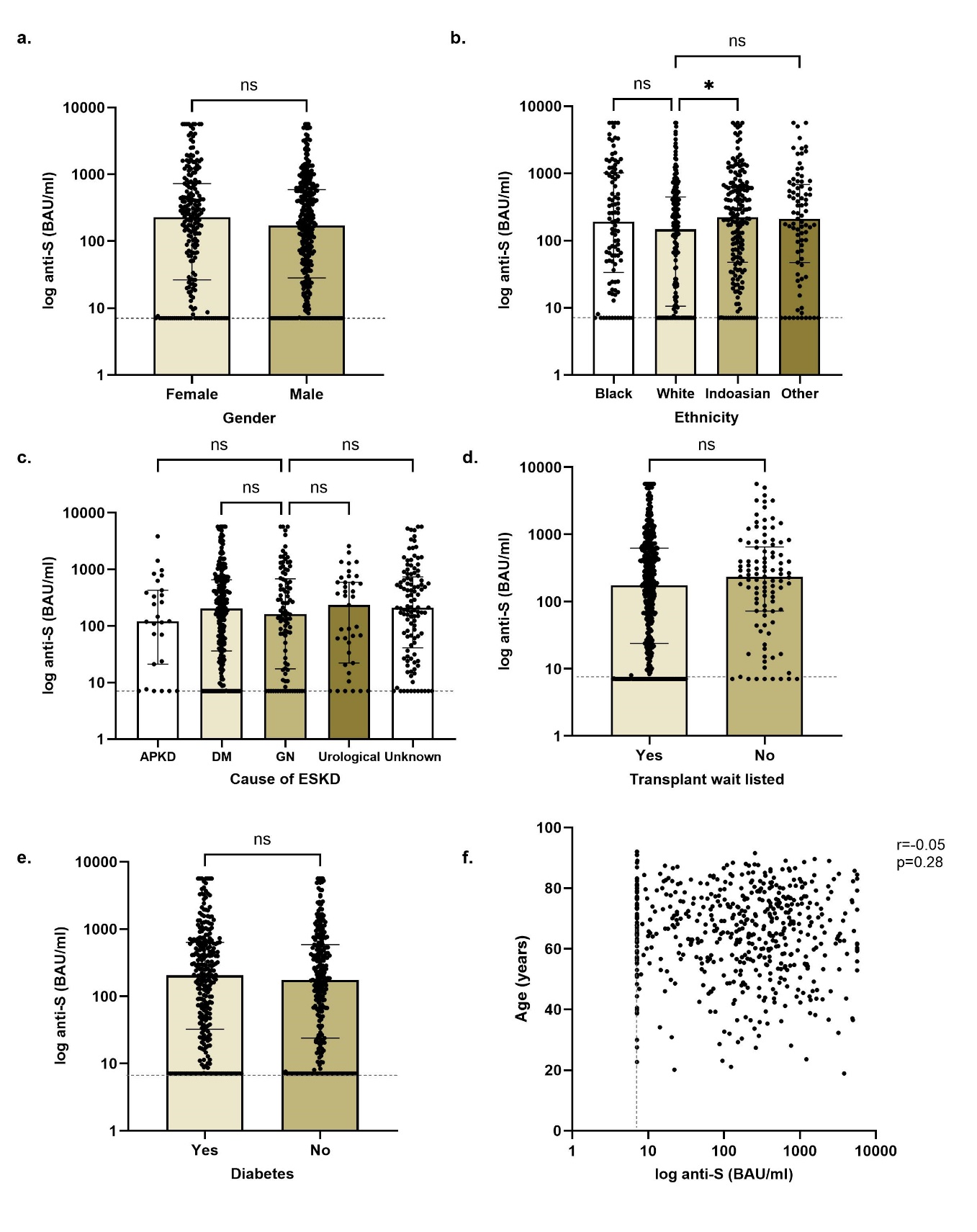

**Table S1. Multivariable analysis of factors associated with a serological response to SARS-CoV-2 vaccination in haemodialysis patients**

| **Variable** | **Reference Group** | **Univariable** | | **Multivariable** | |
| --- | --- | --- | --- | --- | --- |
|  |  | **OR (95% CI)** | **P value** | **OR (95% CI)** | **P value** |
| Age | Years | 0.996 (0.98-1.01) | 0.63 | - |  |
| Ethnicity | White | 0.63 (0.39-1.03) | 0.067 | - |  |
| Immunosuppression | Yes | 0.15 (0.09-0.26) | <0.0001 | 0.15 (0.0.9-0.26) | <0.0001 |
| Vaccine | ChAdOx-1 | 0.67 (0.41-1.09) | 0.11 | - |  |
| Transplant Wait List | Yes | 2.16 (1.00-4.64) | 0.049 | 2.61 (1.16-5.87) | 0.02 |
| Previous Transplant | Yes | 0.36 (0.21-0.61) | 0.0001 | - |  |
| Time at ESKD | Months | 0.997 (0.995-1.00) | 0.019 | - |  |

ESKD end stage kidney disease

**Table S2**. **Characteristics of in patients with prior infection who failed to seroconvert**

| AGE | VACCINE | GENDER | ETHNICITY | MONTHS at ESKD | CAUSE | WAIT LIST | Previous Tranplant | IMMUNO-SUPPRESSION | TYPE IS |
| --- | --- | --- | --- | --- | --- | --- | --- | --- | --- |
| 50-60 | ChAdOx-1 | M | Black | 153 | Unknown | No | No | No | No |
| 60-70 | ChAdOx-1 | M | Black | 6 | Diabetes | No | No | No | No |
| 70-80 | BNT162b2 | M | Indoasian | 142 | Unknown | No | Yes | Yes | CNI |
| >80 | BNT162b2 | F | Black | 24 | Diabetes | No | No | No | No |
| <40 | BNT162b2 | F | White | 15 | Diabetes | No | Yes | Yes | CNI/MMF/Pred |

CNI calcineurin inhibitor; ESKD end stage kidney disease; MMF mycophenolate mofetil

**Figure S2. Comparison of anti-S concentrations by method of diagnosis of previous infection and the presence of co-existing anti-NP**

a. There was no difference in median anti-S in those patients who were diagnosed by PCR compared with serology, with patients receiving BNT162b2 having a median anti-S of 5184 (1773-5680) BAU/ml and 3221 (1379-5680)) BAU/ml respectively, p=0.70, and those receiving ChAdOx-1 median concentrations of 1183 (512-2807) BAU/ml and 2080 (675-3876) BAU/ml respectively, p=0.21

b. The co-existence of anti-NP was associated higher anti-S concentrations, with anti-NP+ and anti-NP- patients receiving BNT162b2 having a median anti-S of 5680 (3338-5680) BAU/ml and 2802 (1097-5680) BAU/ml respectively, p<0.0001; and those receiving ChAdOx-1 2120 (880-4485) BAU/ml and 1472 (636-273) BAU/ml respectively, p=0.01

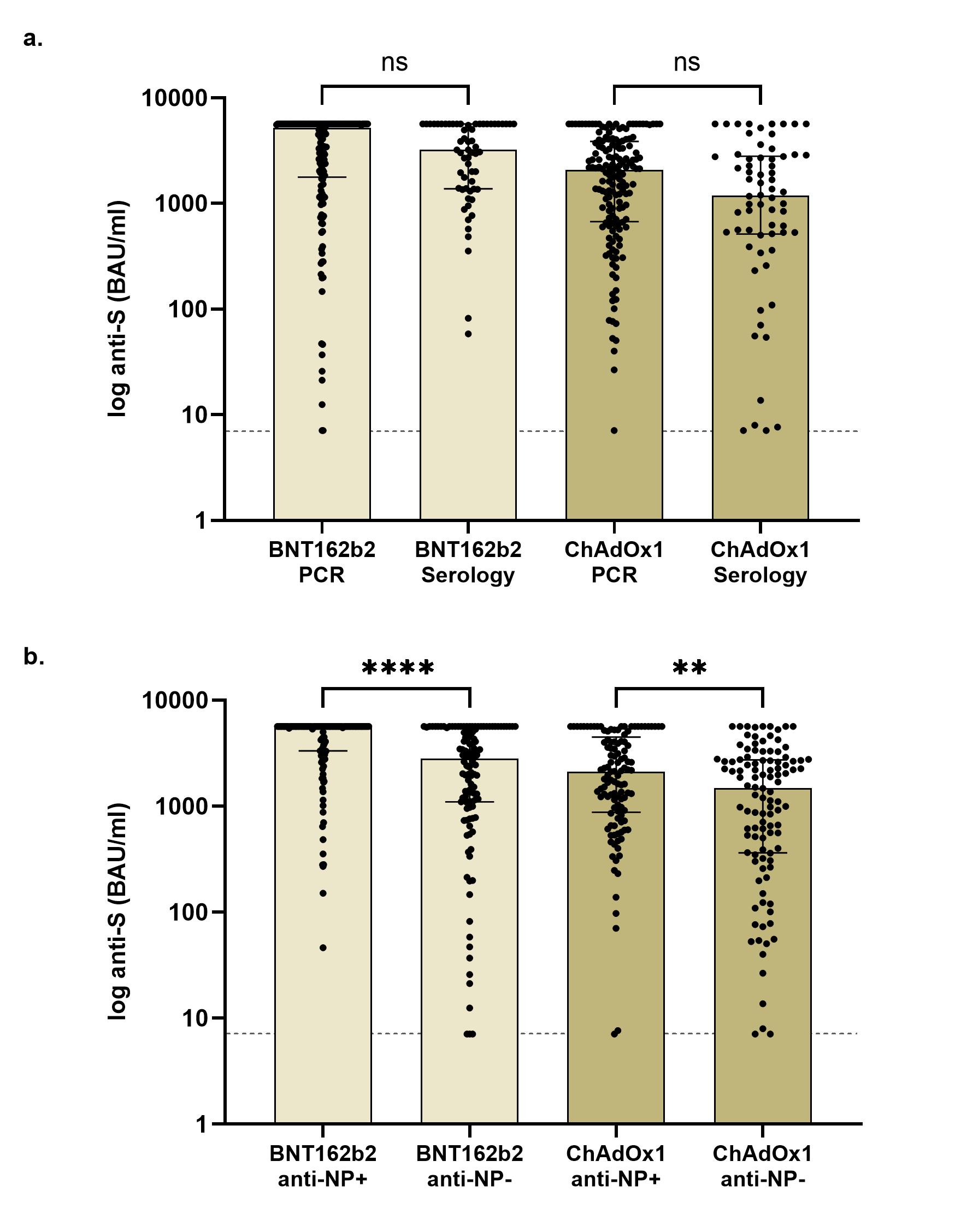

**Table S3**. **Characteristics of 25 patients with no immunological response**

| AGE | VACCINE | GENDER | ETHNICITY | MONTHS at ESKD | CAUSE | WAIT LIST | PRECOVID | IMMUNO-SUPPRESSION | TYPE IMMUNO-SUPPRESSION |
| --- | --- | --- | --- | --- | --- | --- | --- | --- | --- |
| >80 | BNT162b2 | Female | Black | 34 | Unknown | No | No | No |  |
| >80 | BNT162b2 | Female | Black | 92 | Unknown | No | No | No |  |
| 50-60 | BNT162b2 | Female | Black | 13 | Urological | No | No | No |  |
| >80 | BNT162b2 | Female | Black | 24 | Diabetes | No | Yes | No |  |
| >80 | BNT162b2 | Male | Indoasian | 40 | Glomerulonephritis | No | No | Yes | Prednisolone |
| 50-60 | BNT162b2 | Female | Other | 349 | Glomerulonephritis | No | No | Yes | Prednisolone |
| <40 | BNT162b2 | Female | White | 15 | Diabetes | No | Yes | Yes | Pred/CNI/MMF |
| 50-60 | ChAdOx1 | Female | Black | 4 | Diabetes | No | No | No |  |
| 50-60 | ChAdOx1 | Male | White | 2 | Diabetes | No | No | No |  |
| 70-80 | ChAdOx1 | Female | Indoasian | 68 | Diabetes | No | No | No |  |
| 70-80 | ChAdOx1 | Female | Other | 73 | Diabetes | No | No | No |  |
| 70-80 | ChAdOx1 | Male | Other | 19 | Diabetes | No | No | No |  |
| 70-80 | ChAdOx1 | Male | White | 59 | Other | No | No | No |  |
| 60-70 | ChAdOx1 | Male | White | 3 | Other | No | No | No |  |
| 70-80 | ChAdOx1 | Male | Other | 20 | Other | No | No | No |  |
| 70-80 | ChAdOx1 | Female | Other | 129 | Cystic Kidney Disease | No | No | Yes | Pred/CNI |
| 70-80 | ChAdOx1 | Female | Indoasian | 210 | Diabetes | No | No | Yes | CNI |
| 70-80 | ChAdOx1 | Female | White | 163 | Glomerulonephritis | No | No | Yes | Pred/CNI |
| 60-70 | ChAdOx1 | Female | White | 434 | Glomerulonephritis | No | No | Yes | Pred/CNI |
| 40-50 | ChAdOx1 | Male | White | 408 | Glomerulonephritis | No | No | Yes | Pred/CNI |
| 60-70 | ChAdOx1 | Male | Indoasian | 16 | Glomerulonephritis | No | No | Yes | Pred/CNI |
| 40-50 | ChAdOx1 | Male | White | 72 | Other | No | No | Yes | Pred/CNI |
| 60-70 | ChAdOx1 | Male | White | 198 | Diabetes | Yes | No | Yes | CNI |
| <40 | ChAdOx1 | Female | White | 39 | Glomerulonephritis | Yes | No | Yes | Pred/MMF |
| 50-60 | ChAdOx1 | Female | White | 139 | Urological | Yes | No | Yes | CNI |

CNI calcineurin inhibitor; ESKD end stage kidney disease; MMF mycophenolate mofetil
